# Oil and gas well development and coccidioidomycosis risk in California: A case-crossover study

**DOI:** 10.1101/2025.09.19.25336198

**Authors:** Lisa I Couper, David J.X. González, Simon K. Camponuri, Amanda K. Weaver, Gail Sondermeyer Cooksey, Duc Vugia, Seema Jain, John Taylor, John Balmes, Ellen A. Eisen, Justin V. Remais, Jennifer R. Head

## Abstract

**Background:** Coccidioidomycosis is an emerging fungal disease caused by inhaling *Coccidioides* spp. spores. As spores reside in the soil, activities that disturb soil may aerosolize and transport the pathogen, yet the types of activities facilitating transmission remain poorly understood.

**Methods:** We conducted a case-crossover study to estimate the association between exposure to new oil and gas wells (ie, those in preproduction) and risk of coccidioidomycosis among nearby residents. We obtained information on coccidioidomycosis cases reported between 2007 and 2022 in Kern County, California—a county among the top in both oil and gas production and coccidioidomycosis incidence. We compared exposure to preproduction wells within 5 km of each patient residence during “hazard” and “control” periods using conditional logistic regression.

**Findings:** During the study period, 73% of Kern County residents lived within 5 km of at least one preproduction well, and 13% lived within 5 km of ≥23 preproduction wells within a single 90-day period. We estimated that the odds of coccidioidomycosis were 11.0% higher (95% CI: 4.3-18.1%) in the 90 days following exposure to at least one preproduction well within 5 km of the patient residence and that the odds of infection increased by 0.7% (95% CI: 0.4-1.0%) for each additional preproduction well within this distance.

**Interpretation:** We identified a previously unrecognized association between oil and gas development and the transmission of an emerging infectious disease. Given the prevalence of oil and gas development in the study region, its impact on coccidioidomycosis incidence there may be large.

**Research in Context:** *Evidence before the study:* Coccidioidomycosis is an emerging fungal disease caused by the inhalation of airborne *Coccidioides* spores. As these spores reside in the soil, activities that disturb soil—such as construction, farming, earthquakes, and dust storms—have previously been associated with elevated transmission risk. However, the full range of activities that facilitate pathogen transmission remains poorly characterized. Here, we investigated whether oil and gas development contributes to coccidioidomycosis risk as this process involves several soil-disturbing steps (eg, site clearing, leveling, movement of heavy equipment), and occurs at high intensity in an endemic region for the disease. To assess existing evidence, we searched PubMed from database inception to May 27, 2025, for articles published in English using search terms “oil well” OR “gas well” OR “oil and gas construction” OR “oil and gas development” AND “infectious disease” OR “transmission” OR “risk”, and their common textual variants. We identified 31 relevant studies investigating associations between oil and gas development and adverse health outcomes, including preterm birth, low birth weight, cancer diagnoses, upper respiratory symptoms, and all-cause mortality. Only one prior study investigated infectious disease outcomes, finding that high levels of exposure to oil and gas production were associated with moderately elevated COVID-19 severity. We found no prior studies investigating the impact of oil and gas development on the risk of coccidioidomycosis or any other environmental pathogens.

*Added value of the study:* This study identifies a previously unrecognized adverse health outcome of oil and gas development. Using a case-crossover study design focused on Kern County, California—one of the top seven oil-producing counties in the U.S.—we found the development of new oil and gas wells was associated with elevated coccidioidomycosis risk for individuals within five kilometers. Associations were strongest when the well was developed during fall or summer months, when dry soils may be most readily aerosolized. Further, we found that approximately 73% of Kern County residents lived near at least one well developed over the study period (2007-2022), and 13% lived near ≥ 23 wells developed within a single 90-day period. Given this high level of exposure, oil and gas development may be an important driver of transmission in this highly endemic region.

*Implications of all the available evidence:* Our study adds to a growing body of evidence of harmful health impacts of oil and gas development. It also identifies a novel pathway of exposure risk for an emerging fungal disease. As there are currently no vaccines available for coccidioidomycosis and few effective antifungal drugs, identifying and mitigating environmental exposures to fungal spores is critical for protecting public health.

## Introduction

Coccidioidomycosis, or Valley fever, is an emerging infectious disease endemic to the southwestern U.S. It is caused by inhaling spores of *Coccidioides* fungi, which reside in the soil and can become airborne through disturbances such as wind and dust storms, earthquakes, construction, and farming.^1–3^ Infectious spores can travel several miles in dust and be inhaled directly or settle and become resuspended in downwind locations.^4^ While increased exposure to ambient fine mineral dust has been associated with increased incidence of coccidioidomycosis in California,^5^ the specific sources of dust emissions remain largely unidentified.

Oil and gas development has been associated with emissions of mineral dust and several other health-damaging air pollutants, including particulate matter (PM), methane, and volatile organic compounds (VOCs).^6–9^ The construction of new wells may be particularly likely to generate dust as the development process involves many soil-disturbing steps, including transporting heavy equipment, clearing and leveling the site, constructing the well pad, digging water and waste pits, and drilling a bore hole—steps collectively termed ‘preproduction’.^6,10–13^ While several adverse health outcomes have previously been associated with oil and gas development—including preterm birth,^14^ asthma,^15^ reduced lung function,^16^ pediatric hematologic cancer,^17^ COVID-19 severity,^18^ and all-cause mortality^19^—its impact on coccidioidomycosis risk has not been investigated.

Here, we investigated the association between coccidioidomycosis and oil and gas development in Kern County, California—one of the top seven oil-producing counties in the U.S.,^20^ and a region where coccidioidomycosis incidence has increased over six-fold since 2000 (Figures 1, S1).^21^ We conducted a case-crossover study, enabling us to isolate the impacts of exposure to preproduction wells prior to case onset while controlling for individual characteristics such as race, sex, and occupation that may be associated with exposure and/or disease but are mostly time-invariant.^3,22^ We also investigated whether the association between exposure and coccidioidomycosis incidence was modified by patient sociodemographics (age, race/ethnicity, and sex) and/or season, as cases typically peak in the fall, potentially because hot and dry soils promote greater spore aerosolization in this period.^23,24^

**Figure 1.**
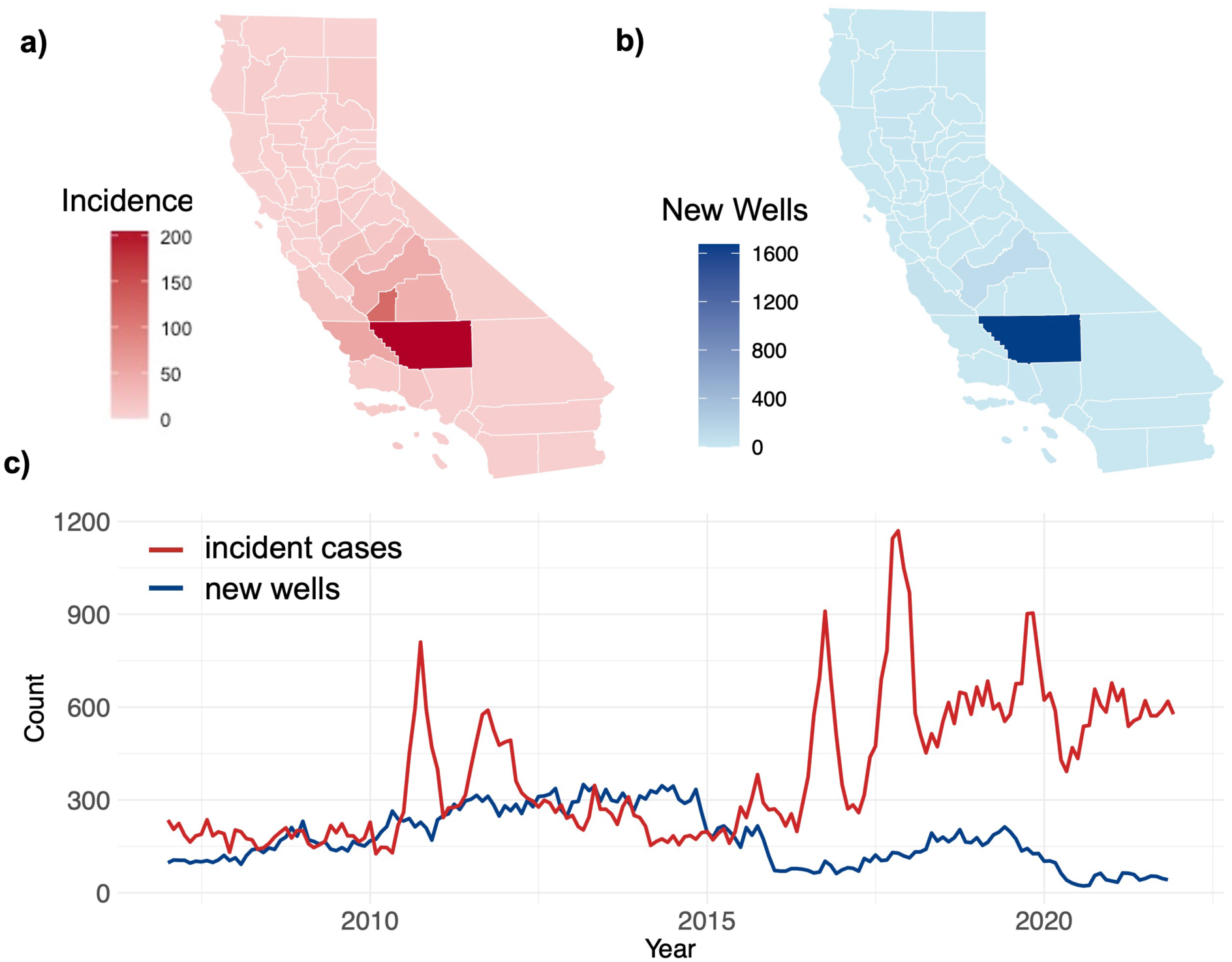
(a) Mean annual county-level coccidioidomycosis incidence (cases per 100,000) in California counties between 2007 and 2022; (b) Mean annual counts of newly constructed wells in California counties between 2007 and 2022; and (c) Monthly counts of newly constructed wells (blue) and cases (red) in Kern County, California.

## Methods

### Study region and period

We restricted our analysis to Kern County, California—the county with the highest incidence of coccidioidomycosis in the U.S. (323.2 per 100,000 in 2018^21^) and the most intensive oil and gas development in the state (Figure 1).^20^ We defined the study period as January 1, 2007 to December 31, 2022—the range for which data on both cases and oil and gas well construction activities were available.

### Case data

We obtained information on all cases of coccidioidomycosis reported in Kern County, California from the California Department of Public Health (appendix p 2). Cases were included in the case-crossover analysis if the patient residence fell within 5 km of a well that was constructed during the study period. This distance threshold was selected given prior evidence of elevated PM_!.#_ up to 5 km from a newly constructed well.^7^ All case data were de-identified prior to analysis. The study received approval from the Committee for Protection of Human Subjects of the California Health and Human Services Agency (CHHS, protocol no. 17-05-2993). Approval by the University of California, Berkeley, was provided by reliance on the CHHS approval.

### Oil and gas well data

We obtained information on all oil and gas wells constructed in California from the California Geologic Energy Management Division (CalGEM) and Enverus, a private data aggregation platform. These data included the geographic coordinates of the well, spud date (when drilling began), completion date (end of the preproduction stage), and the well status (eg, active, idle, cancelled) (Table S1). We subset the data to include only non-cancelled oil and gas wells with a spud and/or completion date during the study period (2007-2022) and located within 5 km of at least one case within the study region. All wells in the dataset had a spud date listed. For wells missing a completion date (85%), we assumed it occurred 14 days after the spud date, following prior work^7^ (but see *Methods: Sensitivity analyses*).

### Exposure definition

We focused our analysis on exposures to wells in preproduction—the period between preparing for spudding and the completion date—to capture potential dust and spore exposures from the steps used to establish and prepare wells for production (Figure S1, Table S1).^7,13,14^ Specifically, we defined the preproduction period for each well as starting one week before the spud date and ending one week after the completion date to account for the full range of oil pad activities that may generate dust, following previous work.^14^ We excluded wells with long gaps between spudding and completion (100+ days; 1.1% of wells), as dust-generating activities associated with preproduction of these wells may have been intermittent or paused.^14^ We classified exposures to wells in preproduction using three metrics: binary (ie, presence or absence of at least one preproduction well), continuous (ie, the number of preproduction wells) and quartile (ie, whether the number of preproduction wells fell within the 1^st^, 2^nd^, 3^rd^, or 4^th^ quartile). For the binary and continuous exposures, we estimated exposures within buffers of varying distances between the patient residence and the well site, ranging from <1 km to 5 km (Figure 2a). For the quartile exposure, we only estimated exposures occurring between 0-5 km from the patient residence. As there were a substantial number of unexposed cases, we calculated quartiles using only non-zero exposures and used unexposed cases as the reference group in statistical models.

**Figure 2.**
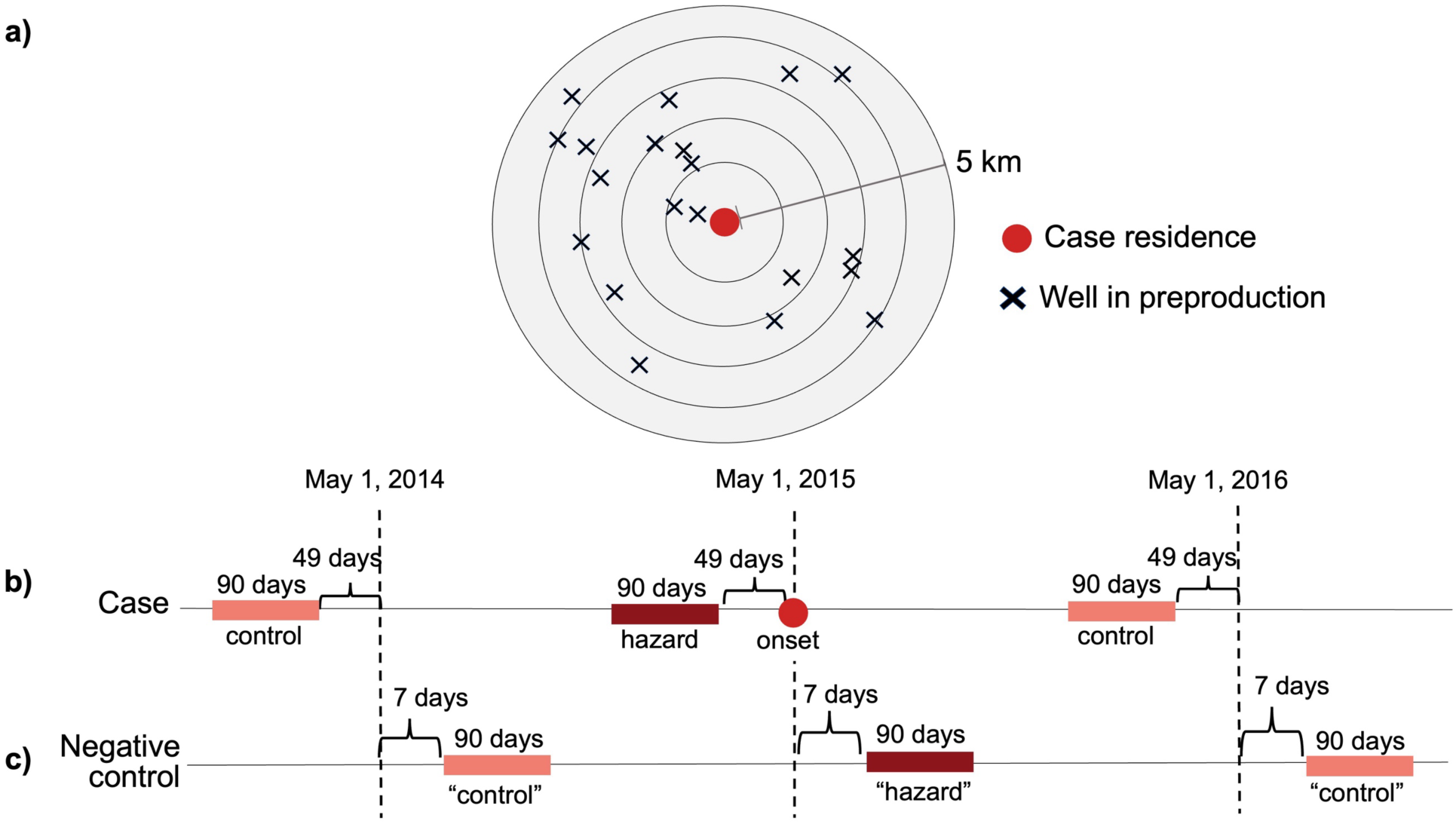
(a) Schematic representing our calculation of exposure to oil and gas wells in preproduction during hazard and control periods (ie, by summing the number of preproduction wells within buffers of varying distances up to 5 km from the patient residence); (b) The case-crossover design, which defined the true hazard period as 49-139 days before case onset, and the control hazard period as the same date range in either the following or preceding year, based on a random draw. See *Methods: Sensitivity analyses* for alternative specifications of the hazard period; and (c) The negative control, which defined the hazard period as the 7-97 days after case onset, and the control hazards as the same date range in the following or preceding year.

### Statistical design

We used a case-crossover approach to examine the association between exposure to preproduction wells and coccidioidomycosis incidence. Herein, inference is obtained by comparing the distribution of exposure during the individual’s hazard period, before the actual outcome occurs, and a corresponding control period (Figure 2). Accordingly, comparisons are made within-individual, allowing each individual to serve as their own control, meaning confounders that are largely time-invariant (eg, sex, race) are inherently adjusted for in the analysis.^22^

We defined the hazard period as the 49-139 days prior to case onset (Figure 2b). Prior work demonstrates that associations between dust exposure and reported coccidioidomycosis incidence are strongest 1-3 months prior to case onset.^5^ This period allows for variability in the time between exposure and symptom onset (estimated at 7-21 days^25,26^), and time between symptom onset and diagnosis (estimated median of 55 days).^27^ To ensure our results are robust to the specification of the hazard period, we repeated the analyses under alternative specifications (see *Methods: Sensitivity analyses*).

For each patient, we selected a control period using a time-stratified semi-symmetric bi-directional design.^28^ Herein, the control period encompassed the same days of the year as the hazard period—to control for seasonality in transmission—but occurred on either the calendar year before or after the hazard period (Figure 2). We used conditional logistic regression to regress case occurrence against exposure during the hazard and control period, using the individual as the strata. We constructed separate models (detailed in appendix p 3) for each exposure metric (binary, continuous, quartile) and, separately, for each of the five exposure distances (0-1, 0-2, 0-3, 0-4, and 0-5 km buffers from the patient residence). Quartile exposures were only assessed over the 0-5 km buffer range.

### Negative controls

While the case-crossover study design, and our approach to control period selection, controls for confounding from time-invariant factors, seasonality, and time trends in the exposure, we sought to further reduce any residual confounding from any unmeasured, time-varying factors by including a negative control exposure in our model, following prior work.^29^ We defined the negative control exposure as a “hazard” period that occurred in the 7-97 days *following* a case report (Figure 2c). This serves as an ideal negative control as the construction of new wells occurring after the estimated disease onset is expected to be associated with the true exposure (as certain locations may experience more frequent oil and gas well construction), yet conditionally independent of disease onset in the absence of confounding, model misspecification, and measurement error.

### Sensitivity analyses

We examined the robustness of our results under alternative specifications of the hazard period and exposure distance. Namely, we repeated the analyses while shifting the length or timing of the hazard period and control periods relative to case onset, or by classifying exposures to wells based on annuli (ie, rings) from the patient residence (appendix p 2).

### Effect Modification

We examined whether the association between the construction of new oil and gas wells and coccidioidomycosis incidence was modified by season and/or patient demographics including age, race/ethnicity (self-reported), and sex, by stratifying models based on these factors (appendix p 2). We used Wald test results to identify differences in the ORs between strata, using a p-value of 0.05 to indicate statistical significance.

### Population exposures to wells in preproduction

To estimate the percent of the population that was exposed to newly constructed wells within 5 km of their residence during the study period, we used population estimates for Kern County from CA-POP, which provides population estimates in 100 m grids based on downscaled census block data (Figure S2).^30^ Using this population estimate, we calculated the percent of the study population exposed to a) at least one well in preproduction during the study period and b) 1-2, 3-7, 8-22, or ≥23 preproduction wells within a single 90-day window in the study period, corresponding to quartiles 1-4. We then identified the highest quartile exposure across 90-day sliding windows of time for each 100 m grid cell (ie, each grid cell fell into a single quartile to avoid double counting).

## Results

### Descriptive statistics

A total of 29,071 coccidioidomycosis cases were reported in Kern County between 2007 and 2022 (Figure 1). During this period, we identified 26,737 newly constructed oil and gas wells (ie, wells in preproduction). To estimate the association between oil and gas well development and coccidioidomycosis incidence, we restricted our analysis to only those cases occurring within 5 km of a well in preproduction anytime during the study period (n = 23,220 cases; 13,890 wells; Table 1, Figure S2). Of these cases, 6,892 (29.7%) had a preproduction well within 5 km of their residence during their hazard period (49-139 days prior to the estimated date of case onset) (Table 2). The number of preproduction wells within 5 km of patient residence during their hazard period varied widely by patient, ranging from 0 to 140 (Table 2).

**Table 1.** Characteristics of the study population: cases in Kern County between 2007 and 2022 that occurred within 5 km of a preproduction well during this period. Characteristics of all Kern County and California residents are included for comparison.

| Characteristic |  | Cases within 5 km of a preproduction well (%)<br>(n = 23,220) | All Kern County residents (%)<br>(n = 916,108) | All California residents (%)<br>(n = 39,040,616) |
| --- | --- | --- | --- | --- |
| Age | 0-17 | 2,568 (11.1%) | 28.4% | 21.8% |
|  | 18-64 | 18,089 (77.9%) | 59.8% | 62.4% |
|  | 65+ | 2,520 (10.9%) | 11.8% | 15.8% |
| Race/<br>Ethnicity | Asian/Pacific Islander | 427 (1.8%) | 6.1% | 16.8% |
|  | Black | 864 (3.7%) | 6.3% | 6.5% |
|  | Hispanic | 6,781 (29.2%) | 56.8% | 40.3% |
|  | Non-Hispanic White | 4,583 (19.7%) | 30.4% | 70.7% |
|  | Unknown | 10,565 (45.5%) |  |  |
| Sex | Female | 10,455 (45.0%) | 48.9% | 49.9% |
|  | Male | 12,715 (54.8%) | 51.1% | 50.1% |
| Season of case onset | Fall | 7,693 (33.1%) |  |  |
|  | Spring | 4,406 (19.0%) |  |  |
|  | Summer | 5,588 (26.1%) |  |  |
|  | Winter | 5,538 (23.9%) |  |  |

**Table 2.** Variation in exposure to wells in preproduction during the hazard period at varying distances from patient residence.

| Distance from patient residence | # cases with non-zero exposure (%) | # preproduction wells exposed to during hazard period (mean ± standard error, range) |
| --- | --- | --- |
| 0-1 km | 116 (0.5%) | 0.02 ± 0.00 (0-25) |
| 0-2 km | 776 (3.3%) | 0.20 ± 0.01 (0-59) |
| 0-3 km | 2,702 (11.6%) | 0.90 ± 0.03 (0-93) |
| 0-4 km | 4,848 (20.9%) | 2.13 ± 0.05 (0-123) |
| 0-5 km | 6,892 (29.7%) | 4.05 ± 0.08 (0-140) |

### Association between the construction of oil and gas wells and coccidioidomycosis

We found significant positive associations between exposure to wells in preproduction and odds of coccidioidomycosis (Figure 3). In particular, in models that classified exposure to preproduction wells as binary, the odds of coccidioidomycosis were 11.0% higher (95% CI: 4.3-18.1%) when there was at least one well in preproduction within 5 km of the patient residence during the hazard period (Figure 3a). Further, in models that represented the number of preproduction wells as continuous, the odds of infection increased by 0.7% (95% CI: 0.4-1.0%) for each additional preproduction well within 5 km of the patient residence (Figure 3b).

**Figure 3.**
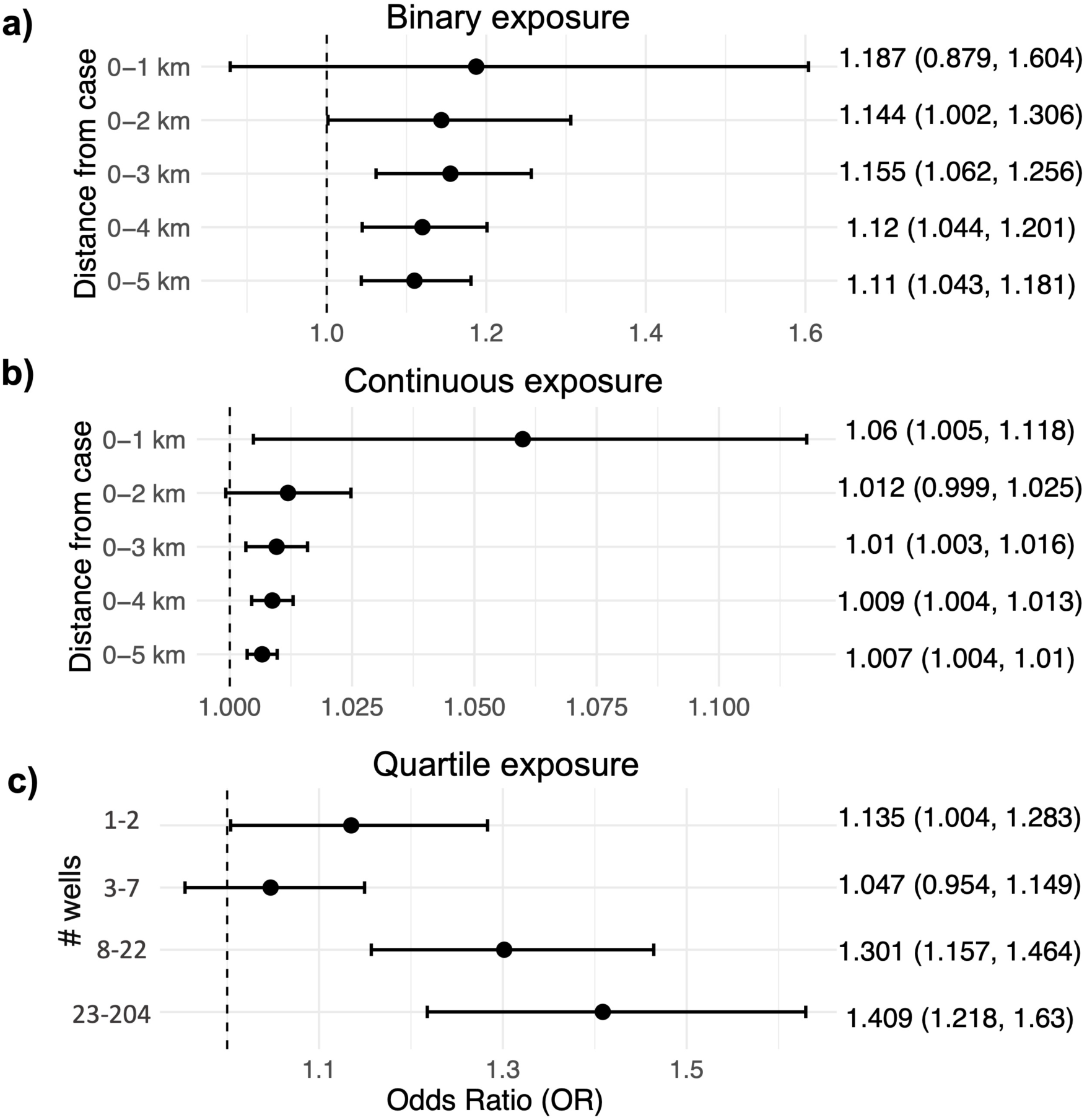
Odds ratios (OR) representing the odds of coccidioidomycosis given exposure to oil and gas well construction at varying distances from the patient residence. The mean and 95% confidence intervals for the ORs are shown for the (a) binary, (b) continuous, and (c) quartile exposure models, and the corresponding values are listed in the right panel. For the quartile exposure, we focus on exposures occurring between 0-5 km and considered only non-zero exposures when calculating quartiles, with the reference group being no exposure to a well in preproduction (see *Methods: Exposure Assessment*). See Figures S4-S6 for the corresponding ORs under alternative specifications.

Similarly, in models that considered quartiles of exposure, odds of coccidioidomycosis were 13.5% higher (95% CI: 0.4-28.3%) given exposure to 1-2 wells (1^st^ quartile) within 5 km of residence compared to 0 wells, and 40.9% higher (95% CI: 21.8-63.0%) given exposure to ≥23 wells (4^th^ quartile) (Figure 3c).

We observed consistent positive associations between oil and gas well construction and risk of coccidioidomycosis for all distance bins. For both binary and continuous exposures, these associations were generally stronger, as indicated by larger point estimates for the ORs, for exposures at closer distances when compared with further distances. However, associations at the 0-1 km distance were not statistically significant under the binary exposure classification, given larger uncertainty in these estimates (ie, wide 95% confidence intervals) (Figure 3a). We may not have had sufficient power to detect associations at this distance as fewer than 1% of cases had exposure to a newly constructed well within 1 km of their residence during the hazard period (Table 2).

For our main model, the ORs for the negative control exposures were typically non-significant, indicating no significant residual confounding, except for the 0-3, 0-4, and 0-5 km distances under the binary exposure model (Figure S3; appendix p 4).

### Sensitivity analyses

Our results were robust to alternative specifications of the hazard period (ie, 21-111, 35-125, 42-132, and 56-146 days prior to case onset as well as the 60-day exposure window) and exposure classification (Figures S4-S7).

### Population exposures

We estimated that approximately 73% of Kern County residents lived within 5 km of at least one well that was in preproduction between 2007 and 2022 (Table 3, Figure S8). Further, approximately 13% were exposed to 23 or more preproduction wells at this distance within a single 90-day period (Table 3, Figure S8).

**Table 3.**
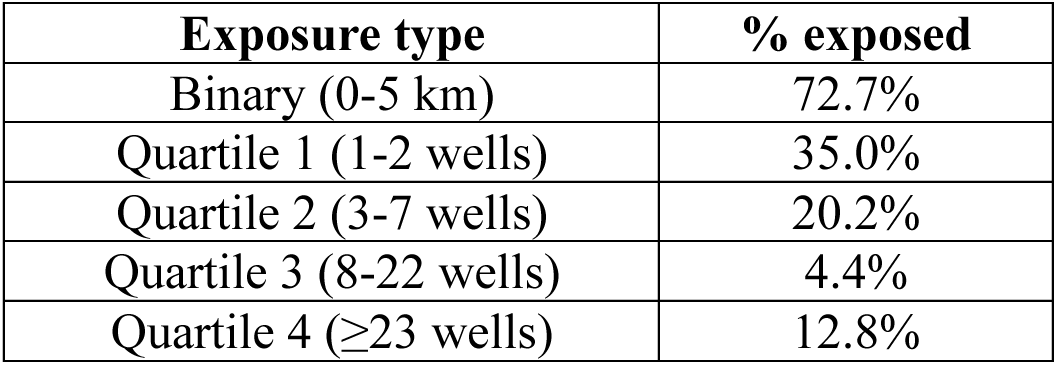
Percent of Kern County residents with varying levels of exposure to wells in preproduction during the study period (2007-2022). The quartile exposures here capture the highest level of exposure within 5 km of their residence for a given individual within a single 90-day period (ie, an individual can only belong to one quartile)

### Effect modification

We observed evidence of effect modification by season of case onset, but not by the patient sociodemographic characteristics we investigated. Specifically, when stratifying analyses by season of case onset, the number of preproduction wells was significantly associated with increased odds of coccidioidomycosis in the fall and winter, but not in other seasons (Figures 4, S9). For each preproduction well within 5 km of the patient residence, the odds of infection in the fall or winter increased by 1.5 and 1.3%, respectively (95% CI: 1.0-2.1% and 0.7-2.0%).

**Figure 4.**
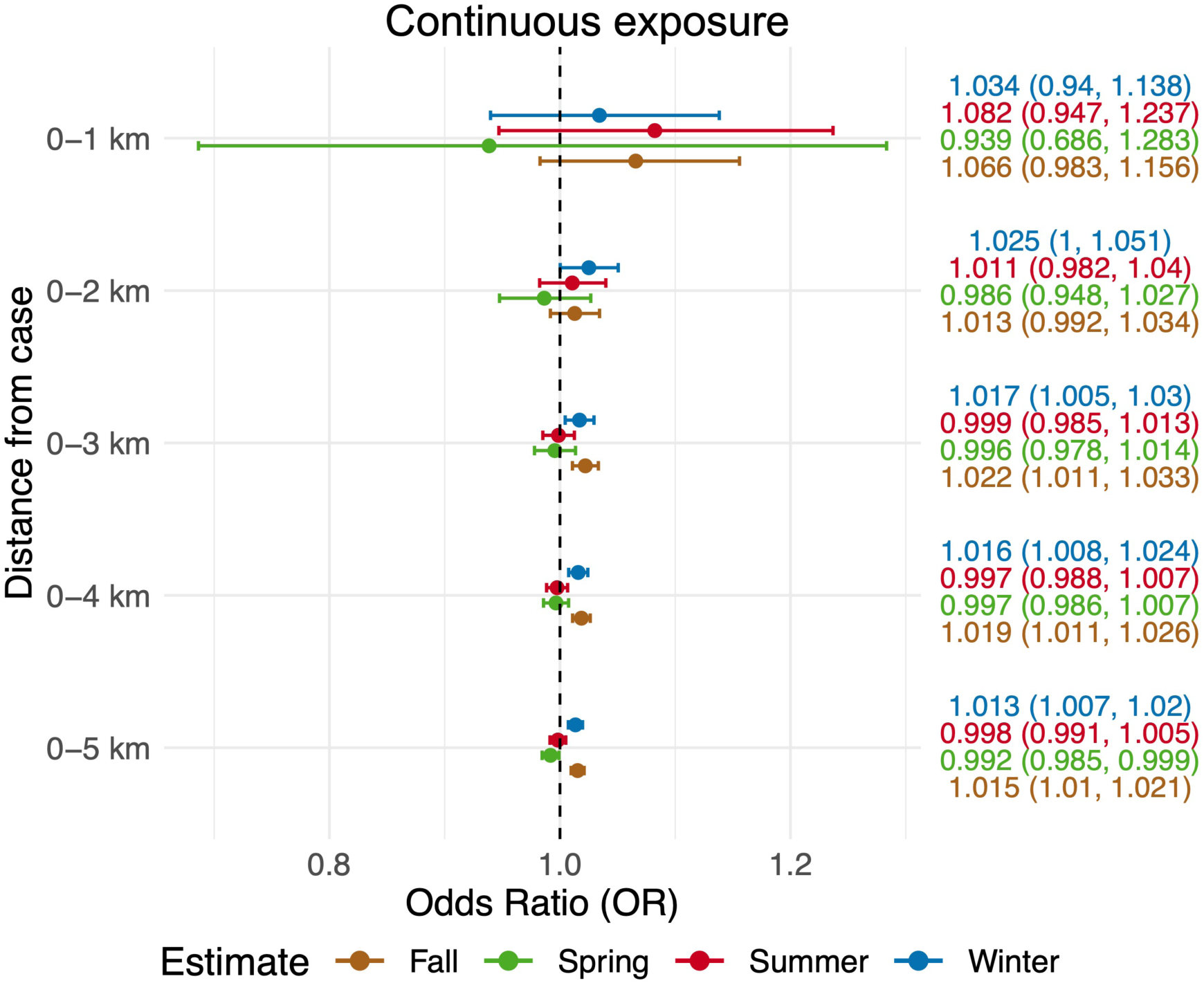
Odds ratios (OR) representing the odds of coccidioidomycosis given exposure to oil and gas well construction at varying distances from the patient residence, stratified by season of case onset. ORs are shown for the continuous exposure model only. See Figure S9 for the season-stratified analyses under the binary exposure model.

Given the lag between exposure window and case onset, this corresponds to wells in preproduction during the summer and fall, respectively. We did not detect consistent evidence of effect modification by patient age, race/ethnicity (self-reported), or sex (Figures S10-S12; appendix p 4).

## Discussion

We identified a previously unrecognized association between oil and gas development and an emerging infectious disease. We found that the construction of new oil and gas wells—a process known to involve soil disturbance^6,7^—was significantly associated with increased risk of coccidioidomycosis in a highly endemic region (Kern County, California). Specifically, the odds of infection were approximately 11% higher given at least one newly constructed (‘preproduction’) well within five kilometers of a patient residence in the months preceding case onset (Figure 3a) and increased by approximately 0.7% for each additional preproduction well at this distance. We observed larger increases in risk given exposure to closer wells (<1 km compared to <5 km), though associations at closer distances were not always statistically significant, potentially due to small sample sizes, as fewer than 1% of cases had any preproduction wells within one kilometer prior to disease onset (Table 2).

Conversely, exposure to preproduction wells at further distances was common: over 70% of Kern County residents had a preproduction well within five kilometers of their residence during the study period (2007-2022) and roughly 13% had 23 or more preproduction wells within five kilometers of their residence in a single three-month period (Table 3). Notably, the five-kilometer threshold used here was motivated by prior work finding elevated PM_!.#_ from newly constructed wells was detectable up to this distance.^7^ The high level of exposure amongst this population, and increased risk of infection given exposure, suggest a potentially large population-level impact of oil and gas well construction on coccidioidomycosis incidence in this region. Population exposure may have decreased in recent years as the number of newly constructed wells was highest during the 2010-2020 period and declined thereafter.

We found that the association between oil and gas well construction and coccidioidomycosis varied by season of case onset, but not by the patient sociodemographics we investigated. In particular, associations were stronger for cases with onset in fall and winter, most closely reflecting exposure to wells in preproduction during the summer and fall (Figure 4). This finding aligns with prior evidence that exposure to dust during the summer and fall months poses a higher risk for coccidioidomycosis compared to other seasons, putatively due to hot and dry conditions promoting greater spore lysis and airborne dispersal.^23,24^

Although several prior studies have found associations between coccidioidomycosis severity and older age, male sex, and racial heritage (namely African and Filipino), we did not observe consistent effect modification by age, sex, or self-reported racial/ethnic identity (Figures S10-S12). This may be because our study design investigates variation in coccidioidomycosis onset given exposure, while prior studies often capture variation in both exposure and disease severity. For example, associations between male sex and coccidioidomycosis hospitalization^31^ may reflect greater occupational exposure to *Coccidioides* (eg, through construction or agricultural work).^32,33^ As our study did not assess variation in pathogen concentration or case severity, further work is needed to disentangle whether and how demographic characteristics may be risk factors for *Coccidioides* exposure versus disease outcomes.

The observed associations between oil and gas well construction and the risk of coccidioidomycosis were established while rigorously controlling for time invariant and time-varying confounders. In particular, our use of a case-crossover study design enabled us to isolate the impacts of exposure to preproduction wells while controlling for time-invariant individual characteristics that may be associated with exposure and/or disease such as race and sex, as well as mostly time-invariant characteristics such as occupation and socioeconomic status.^3,22^ Our selection of control periods following a semi-symmetric bi-directional design that matches control period to the same time of year as the hazard period further controls for confounding by seasonality or time trends in the exposure. Our use of negative control exposures in models (here, exposure to preproduction wells that occurred after case onset, which thus could not cause the case) helps adjust for residual confounding due to unmeasured time-varying characteristics.^29^

Our study has several limitations, including the potential for exposure misclassification. We assessed exposure based on reported residence at the time of diagnosis, which may not reflect the true location of exposure to *Coccidioides* spores (eg, workplace exposure). Similarly, we assigned case date based on the patients’ estimated symptom onset date, and, when unavailable (92% of cases), we used the earliest recorded clinical or laboratory date. As the incubation period, time to health-care seeking, and diagnostic testing can vary between individuals, we considered a three-month exposure period (1.5-4.5 months prior to case onset) to account for this variation, and conducted sensitivity analyses applying different lags between exposure and case onset. However, if the times between exposure and case onset were substantially different, our effect estimates may also be subject to exposure misclassification. In both instances, the potential exposure misclassification would likely be nondifferential between the hazard and control periods, thereby biasing our associations towards the null. Further, our sensitivity analyses confirmed that our results were robust to alternative specifications of lags between exposure and case onset. Lastly, we only had data on reported cases, and therefore did not capture individuals with coccidioidomycosis who did not experience symptoms, did not seek medical care, or were misdiagnosed. This may have reduced our power to detect associations between oil and gas production and infection risk and could have biased our associations towards the null if individuals living near oil and gas wells and other hazards are less likely to seek medical care.

Despite these limitations, our study adds to a growing body of evidence of harmful health impacts of oil and gas development. In particular, prior research in California has found associations between oil and gas development and adverse birth outcomes,^14^ asthma,^15^ and lower lung function.^16^ Here, we identify a previously unrecognized health consequence of oil and gas development—transmission of an emerging infectious disease—likely directly related to the disturbance of dirt and soil during well construction. As there are currently no vaccines available for coccidioidomycosis, limited therapeutic options, and rising concerns of antifungal resistance,^26^ measures aimed at reducing environmental exposure to *Coccidioides* spores are critical to protecting public health. There are numerous additional risk factors for coccidioidomycosis relevant in this region including agricultural or construction work, excavation of undisturbed land unrelated to oil and gas drilling, earthquakes, and dust storms.^32–35^ These risk factors are not mutually exclusive with exposure to oil and gas well construction, meaning that this study population could be exposed to *Coccidioides* via multiple sources.

Measures including wetting soil prior to disturbance to reduce dust, avoiding dusty areas when possible, and, when unavoidable, keeping windows closed and/or wearing a properly-fitting N95 respirator can all help reduce risk of infection.^36^ People living in endemic regions near sites where major soil disturbance activities are planned should be made aware of the potential risk in order to prepare for personal prevention measures and to seek care and possible coccidioidomycosis testing in a timely manner if compatible symptoms develop. Further, our results suggest that shifting soil-disrupting activities away from summer and fall months could further help avoid coccidioidomycosis cases. The decreasing number of newly constructed wells in this region in recent years may decrease the associated risk for coccidioidomycosis.

Additional research is needed to clarify how risk factors for *Coccidioides* exposure (eg soil-disturbing occupations or recreational activities, proximity to dust-generating events) interact and the extent to which intervening on these factors would reduce risks. Further work is also needed to understand the mechanistic links between the construction of oil and gas wells and infection, including identifying the types of activities most likely to aerosolize spores, the distance over which aerosolized spores may travel, and the role of wind in their dispersal.

## Data sharing

The R script used to conduct the data analysis is available in the publicly available GitHub repository: https://github.com/lcouper/OilWellsAndCocci. Human case data are protected health information and can only be obtained by submitting a formal data use request to the California Department of Public Health (CDPH), Infectious Disease Branch, Surveillance and Statistics Section. Oil and gas data are publicly available from the California Geologic Energy Management Division (CalGEM) or through purchase from Enverus, a private data aggregation platform.

