## appendix for "Oil and gas well development and coccidioidomycosis risk in California: A case-crossover study"


**Supplementary Methods**
Case data
Sensitivity analyses
Effect modification
Statistical design

**Supplementary Results**
Sensitivity analyses
Effect modification
Negative controls

**Supplementary Table S1**
**Table S1.** Timeline of activities associated with oil and gas development

**Supplementary Figures S1 - S12**
**Fig S1.** DAG representing hypothesized relationships between well construction and coccidioidomycosis risk
**Fig S2.** Population size estimates and location of newly constructed wells used in analysis
**Fig S3.** OR and negative controls
**Fig S4.** OR estimated under alternative model specifications (binary exposure)
**Fig S5.** OR estimated under alternative model specifications (continuous exposure)
**Fig S6**. OR estimated under alternative model specifications (quartile exposure)
**Fig S7**. OR estimated using annuli from patient residence
**Fig S8.** Percent of cases with varying levels of well exposure at each distance
**Fig S9.** OR stratified by season (binary exposure)
**Fig S10.** OR stratified by age
**Fig S11**. OR stratified by sex
**Fig S12.** OR stratified by race/ethnicity

**Supplementary References**

**Supplementary Methods**

*Case data*

We obtained information on all cases of coccidioidomycosis reported in Kern County, California from the California Department of Public Health. Coccidioidomycosis has been reportable in California since 1995. Since 2019, patients are classified as coccidioidomycosis cases based on a positive *Coccidioides* laboratory test, while prior to 2019 classification required both laboratory and clinical criteria to be met.[1] For a given patient, cases are only recorded once, thus patients with more than one reported positive *Coccidioides* test are only recorded upon their initial positive result. When possible, the date of onset was recorded for each case from patient interview. However, this was only available for 8% of cases in our study population and period. For all other cases, the earliest recorded clinical or laboratory date was used as the estimated onset date. Thus, the estimated date of onset reflects delays in spore inhalation and symptom onset, diagnosis, specimen collection, record creation, case reporting, and/or patient death. Prior work has found that, for coccidioidomycosis patients who could recall their estimated date of disease onset, the median time between symptom onset to diagnosis date was 55 days.[2] As the estimated location of exposure to *Coccidioides* is not available in the surveillance data, case locations were assigned to the latitude and longitude of the patient residence. Ninety-four percent of cases in our study population and period could be successfully geocoded to a residential address.

*Sensitivity analyses*

To assess the robustness of our results, we repeated the analyses under alternative specifications of the hazard period and exposure distance. Specifically, we repeated the analyses while shifting the hazard period from 49-139 days prior to case onset (as in the main model) to 21-111, 35-125, 42-132, and 56-146 days prior, to account for varying delays between transmission and estimated onset as well as pre-spudding activities that may generate dust. We also repeated the analysis using a shorter hazard period (60 days, compared to 90 days in the main model) to examine model robustness under a narrower exposure window. For each shift of the hazard period, we updated the control period accordingly to retain the match by calendar year.

To assess the sensitivity of our results to assumptions concerning the relative timing of spud and completion dates, we repeated the analysis while focusing on exposures to well spudding alone rather than the full preproduction period. Herein, we defined the exposure interval for each well as the 7 days prior to and including the spud date.

Lastly, we repeated the analysis while classifying exposure to wells in 1 km annuli (ie, rings) away from the patient residence (ie, 0-1, 1-2, 2-3, 3-4, and 4-5 km). Here, we constructed a single multivariate conditional logistic regression model, including five exposure variables representing either the presence or absence of preproduction wells (binary exposure) or the number of preproduction wells (continuous exposure) in each annulus. We found the strongest positive associations at the 4-5 km distance in the continuous exposure model, and non-significant associations at most other annuli (Figure S7).

*Effect modification*

We examined whether the association between the construction of new oil and gas wells and coccidioidomycosis incidence was modified by season and/or patient demographics including age, race/ethnicity, and sex. To do so, we stratified models by season of case onset (winter: December-February, spring: March-May, summer: June-August, fall: September-November). Similarly, we stratified models by patient characteristics using the following strata: age group (0-17, 18-64, and 65+ years of age), race/ethnicity (Asian/Pacific Islander, Black, Hispanic, and non-Hispanic White), and sex (male, female). Information on race/ethnicity was self-reported, and available for 58.0% of patients in our study population and period. We note that self-reported race/ethnicity may differ from genetic ancestry, and that there is no scientific definition for the groupings used here.

*Statistical design*

We used a case-crossover approach to examine the association between exposure to preproduction wells and coccidioidomycosis incidence. Inference is obtained by comparing the distribution of exposures during the individual's hazard period, before the actual outcome occurs, and a corresponding control period. For each patient, we selected a control period using a time-stratified semi-symmetric bi-directional design.[3] Herein, the control period encompassed the same days of the year as the hazard period—to control for seasonality in transmission—but occurred on either the calendar year before or after the hazard period, with the directionality based on a random draw. This semi-symmetric bi-directional design has been shown to minimize confounding due to time trends in the exposure, under the assumption that the occurrence of the outcome does not affect the probability of subsequent exposure. That is, we assume that having coccidioidomycosis does not affect the future probability of exposure to new wells.[3,4]

We used conditional logistic regression to regress case occurrence against exposure during the hazard and control period, using the individual as the strata. We constructed separate models for each exposure metric (binary, continuous, quartile) and, separately, for each of the five exposure distances (0-1, 0-2, 0-3, 0-4, and 0-5 km buffers from the patient residence). Quartile exposures were only assessed over the 0-5 km buffer range. Models were of the following form:

$$logit(Y_{ip}) = \alpha_i + \beta_1 \times Exposure_{i,p,metric,dist} + \beta_2 \times Negative\ Control\ Exposure_{i,p,metric,dist} \quad [1]$$

where $Y_{ip}$ is a binary indicator for case status of patient $i$ during period $p$ (taking the value of 1 during the hazard period and 0 during the control period), $\alpha_i$ represents the patient-specific intercept obtained from using the individual as the strata, and $Exposure_{i,p,metric,dist}$ represents the exposure of interest for patient $i$ during period $p$ (hazard or control period), calculated for a given metric (binary, continuous, quartile) and within a given exposure distance. When exponentiated, the model coefficient on the true exposure, $\beta_1$, represents the odds ratio, or the relative change in odds of coccidioidomycosis incidence associated with a one unit increase in the exposure (herein, presence vs absence, one additional well, or one quartile increase).

Models were implemented in R version 4.3 using the clogit function in the *survival* package.[5]

**Supplemental Results**

*Sensitivity analyses*

When shifting the hazard and control periods relative to case onset or using a shorter hazard and control period, we found the associations between exposure to oil and gas development and odds of coccidioidomycosis were slightly attenuated compared to those of the main model, but remained statistically significant in most instances (Figures S4-S6). When classifying exposure to wells in 1 km annuli away from the patient residence, we found the strongest positive associations at the 4-5 km distance in the continuous exposure model, and non-significant associations at most other annuli (Figure S7).

*Effect modification*

We detected evidence of effect modification by season of case onset, but no consistent evidence of effect modification by patient age, race/ethnicity, or sex (Figures S10-S12). That is, the 95% confidence intervals of the ORs overlapped and/or the Wald test result was non-significant ($p > 0.05$) for nearly all strata in each distance bin and exposure model with two exceptions. First, we observed a stronger positive association between the presence of preproduction wells within 4 km of patient residence and risk of infection for Asian/Pacific Islander (AAPI) individuals relative to Black, Non-Hispanic White, and Hispanic individuals ($p < 0.04$ for all, Figure S12). We also observed a stronger positive association between the number of preproduction wells within 3 km and risk of infection for the 0-17 age category relative to the 18-64 and 65+ categories ($p < 0.05$ for both groups, Figure S10). However, in both cases, the effect modification was only observed at a single distance and exposure classification. Further, the AAPI race/ethnicity category and 0-17 age category constituted a relatively small portion of the overall study population (1.8% and 11.1%, respectively; Table 1), thus these results could be spurious due to the small sample sizes.

*Negative controls*

We sought to adjust for any residual individual-level confounding in our analysis by including a negative control exposure in our model. For our main model, the ORs for the negative control exposures were typically non-significant, indicating no residual confounding, except for the 0-3, 0-4, and 0-5 km distances under the binary exposure model (Figure S3). The positive ORs on the negative controls observed in these scenarios suggest that the presence of a preproduction well at these distances in the hazard and control period may be correlated (ie, specific locations may experience frequent oil and gas well construction). However, inclusion of the negative control in each model helps control for any variation due to residual confounding, thereby minimizing bias due to confounding in our results.[6,7]

**Table S1.** Timeline of activities commonly associated with oil and gas development. We note this list of activities may be non-exhaustive.

| **Time Period** | **Associated activities** |
|---|---|
| 1-week pre-spudding | Road construction, equipment transport, site clearing and leveling, digging of pits for water and waste |
| Preproduction (spud date to completion date) | Preparation of the well pad, road construction, initial drilling ('spudding'), and well completion |
| 1-week post-completion | Truck traffic, site maintenance |
| Production | Oil and gas extraction (eg, cyclic steam injection, water/steam flooding, hydraulic fracturing), fugitive emissions from equipment |

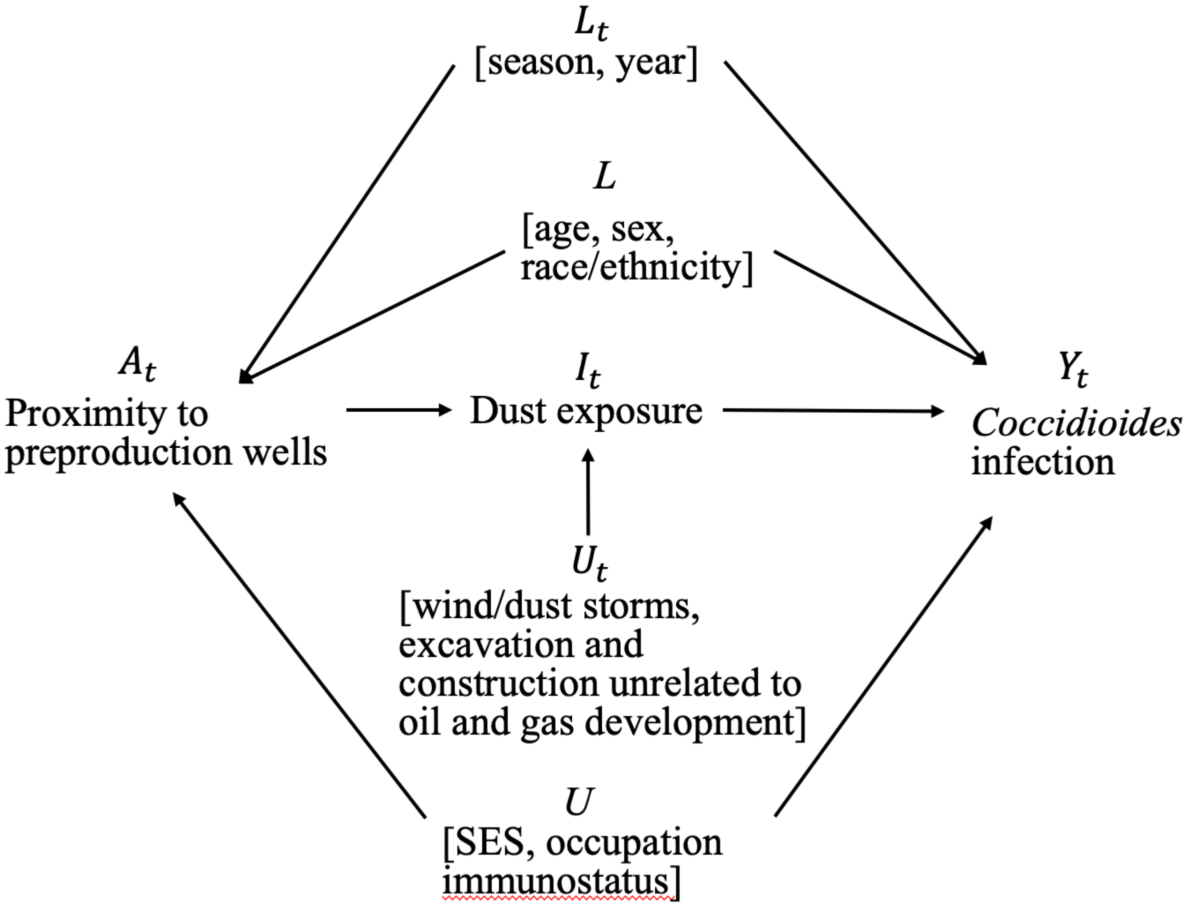


**Figure S1.** Directed acyclic graph (DAG) representing the hypothesized relationships between proximity to oil and gas wells in preproduction ($A_t$) and *Coccidioides* infection ($Y_t$), including measured time-varying ($L_t$) and time-invariant ($L$) confounders, unmeasured time-varying ($U_t$) and time-invariant ($U$) confounders, and intermediate factors ($I_t$). Here, 'measured' indicates that data on these factors were available. SES denotes socioeconomic status.

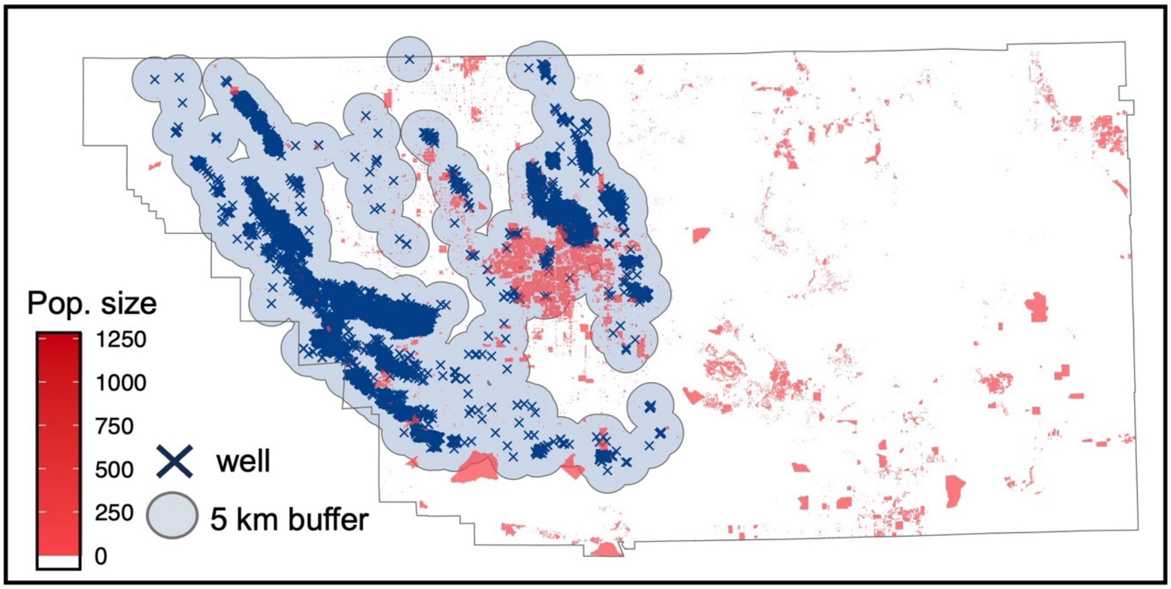


**Figure S2.** Population size estimates and location of newly constructed wells used to estimate the percent of Kern County residents exposed to oil and gas well construction (see *Methods: Population exposures*). Population estimates are provided in 100 m grid cells for 2020 from CA-POP. Non-colored grid cells denote un-populated regions.

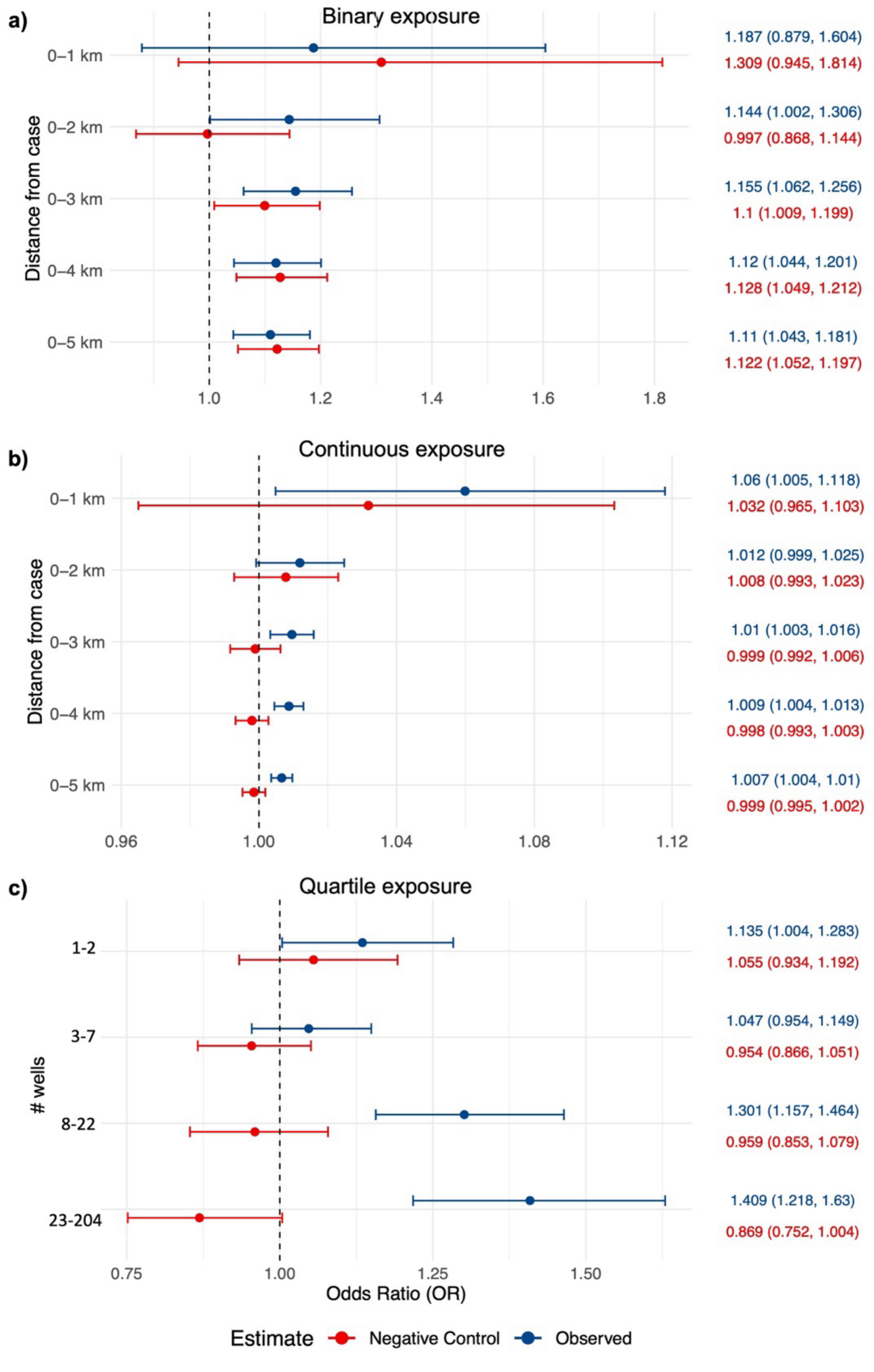


**Figure S3.** Odds ratios (OR) representing the odds of coccidioidomycosis given exposure to oil and gas well construction at varying distances from the patient residence for the observed (red) and negative control (blue). Odds ratios are shown for the (a) binary, (b) continuous, and (c) quartile exposure models. For the quartile exposure, we focus on exposures occurring between 0-5 km and considered only non-zero exposures when calculating quartiles (see *Methods: Exposure Assessment*).

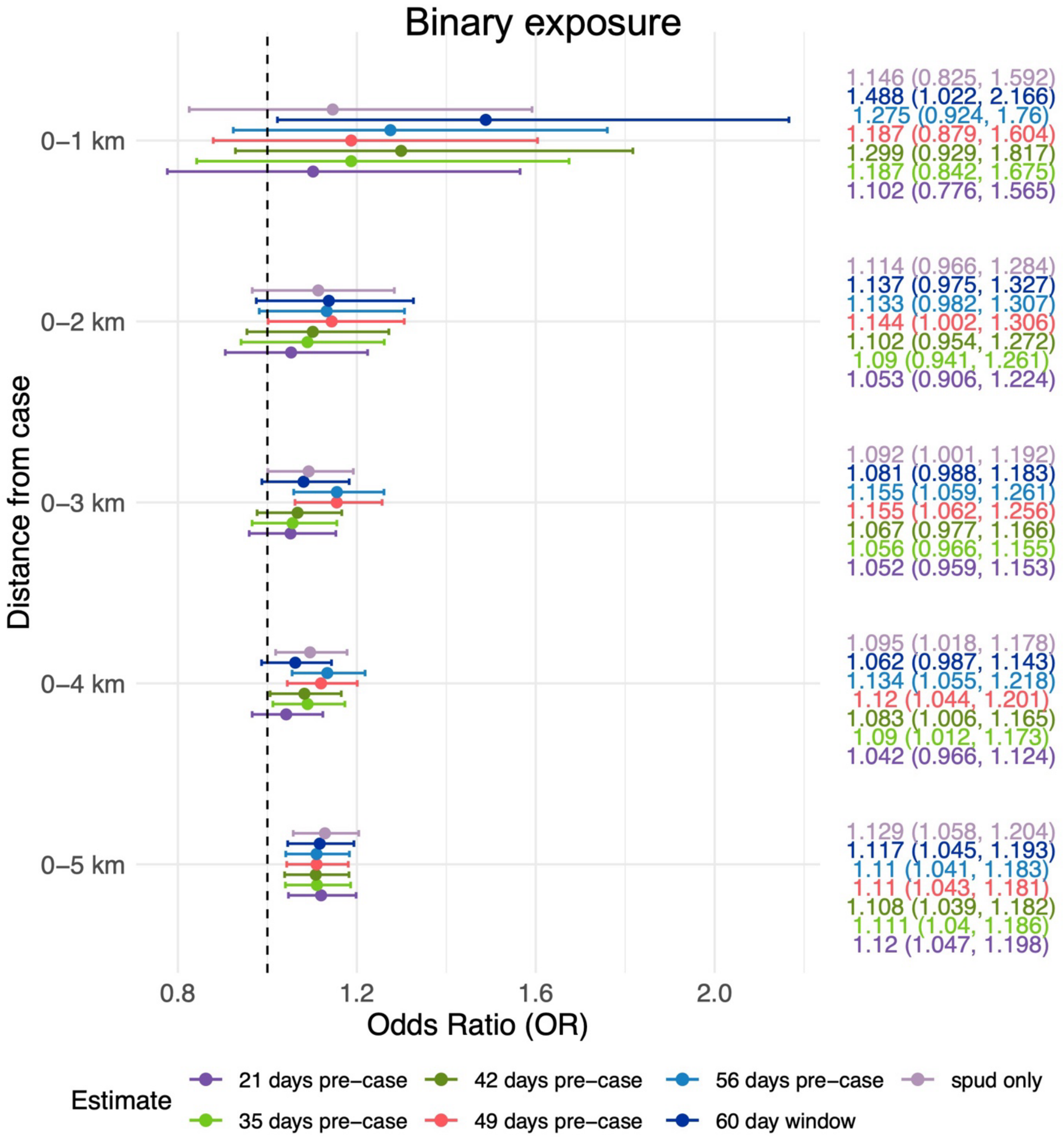


**Figure S4.** Odds ratios (OR) representing the odds of coccidioidomycosis given exposure to oil and gas well construction at varying distances from the patient residence under alternative model specifications. These include alternative specifications of the hazard period: 21-111, 35-125, 42-132, 49-139 (main model), and 56-146 days prior to case onset; the length of the exposure window (60 days, compared to 90 days as used in the main model and all other specifications); and exposures focused on spudding alone rather than the full preproduction period. Odds ratios are shown for the binary exposure models. See Figures S5 and S6 for ORs from the continuous and quartile models.

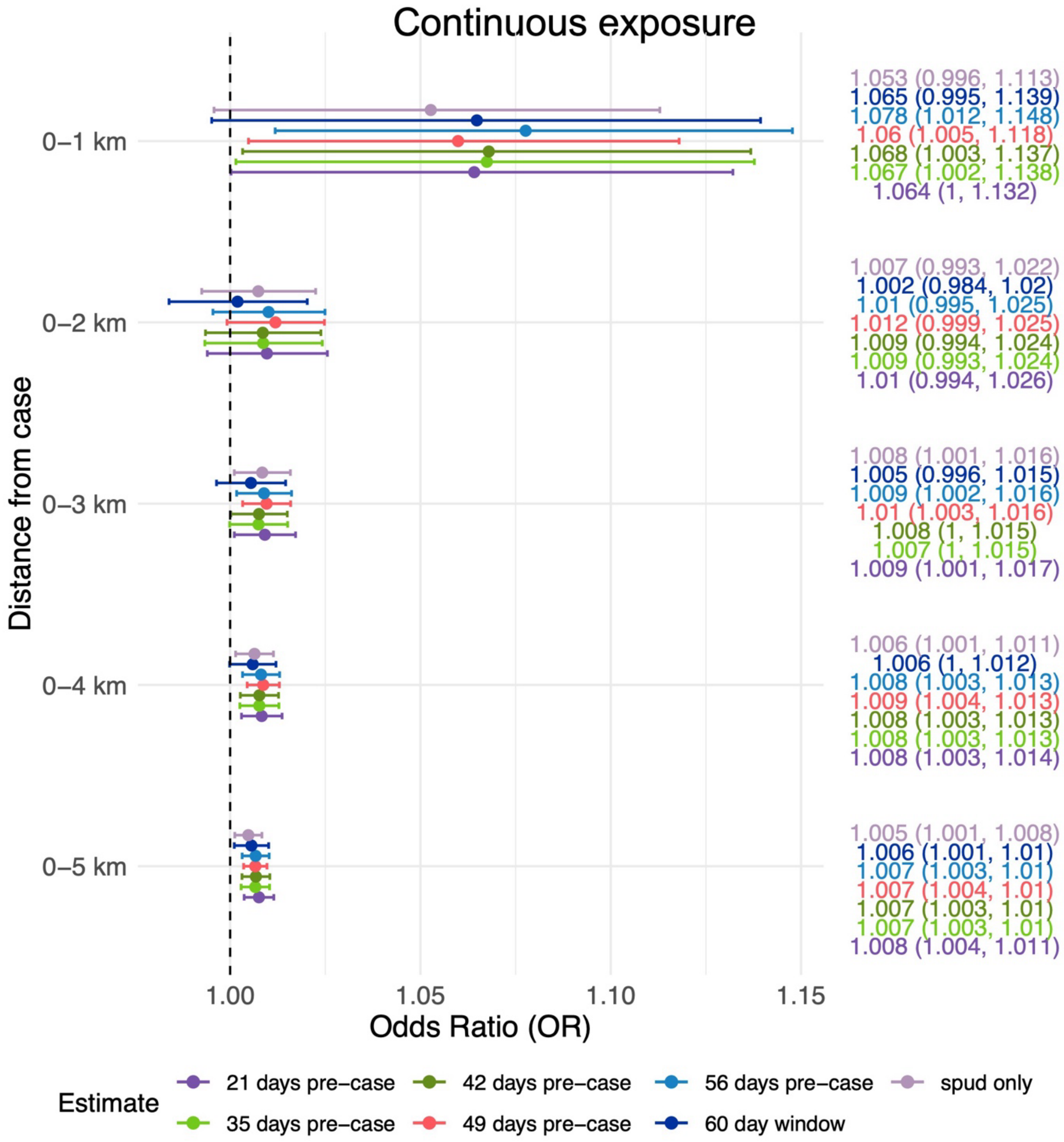


**Figure S5.** Odds ratios (OR) representing the odds of coccidioidomycosis given exposure to oil and gas well construction at varying distances from the patient residence under alternative model specifications. These include alternative specifications of the hazard period: 21-111, 35-125, 42-132, 49-139 (main model), and 56-146 days prior to case onset; the length of the exposure window (60 days, compared to 90 days as used in the main model and all other specifications); and exposures focused on spudding alone rather than the full preproduction period. Odds ratios are shown for the continuous exposure models. See Figures S4 and S6 for ORs from the binary and quartile models.

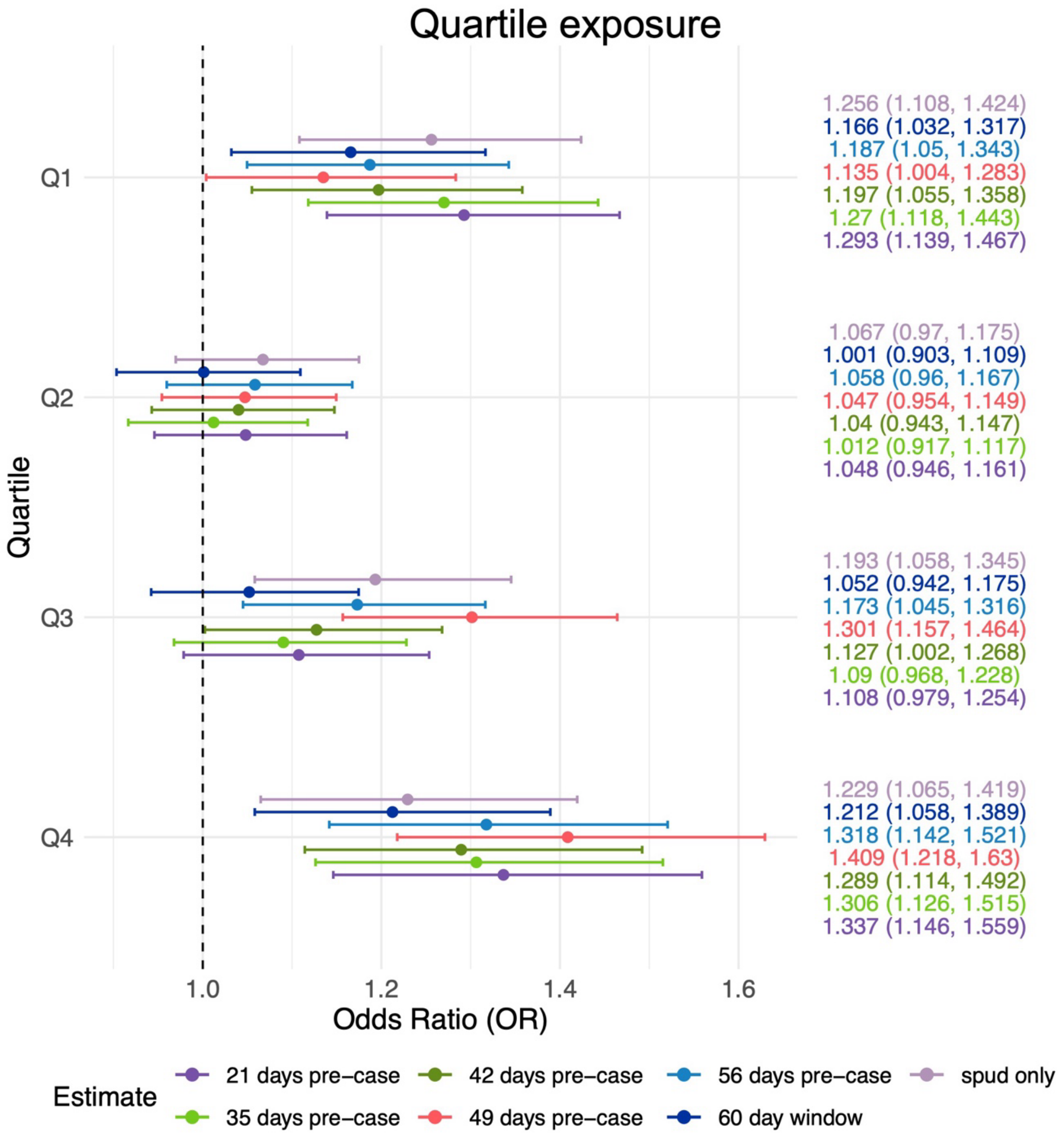


**Figure S6.** Odds ratios (OR) representing the odds of coccidioidomycosis given exposure to oil and gas well construction at varying distances from the patient residence under alternative model specifications. These include alternative specifications of the hazard period: 21-111, 35-25, 42-132, 49-139 (main model), and 56-146 days prior to case onset; the length of the exposure window (60 days, compared to 90 days as used in the main model and all other specifications); and exposures focused on spudding alone rather than the full preproduction period. Odds ratios are shown for the quartile models, which focuses on exposures occurring between 0-5 km and considered only non-zero exposures (see *Methods: Exposure Assessment*). Note that the values of the quartiles differ slightly between hazard specifications. See Figures S4 and S5 for ORs from the binary and continuous models.

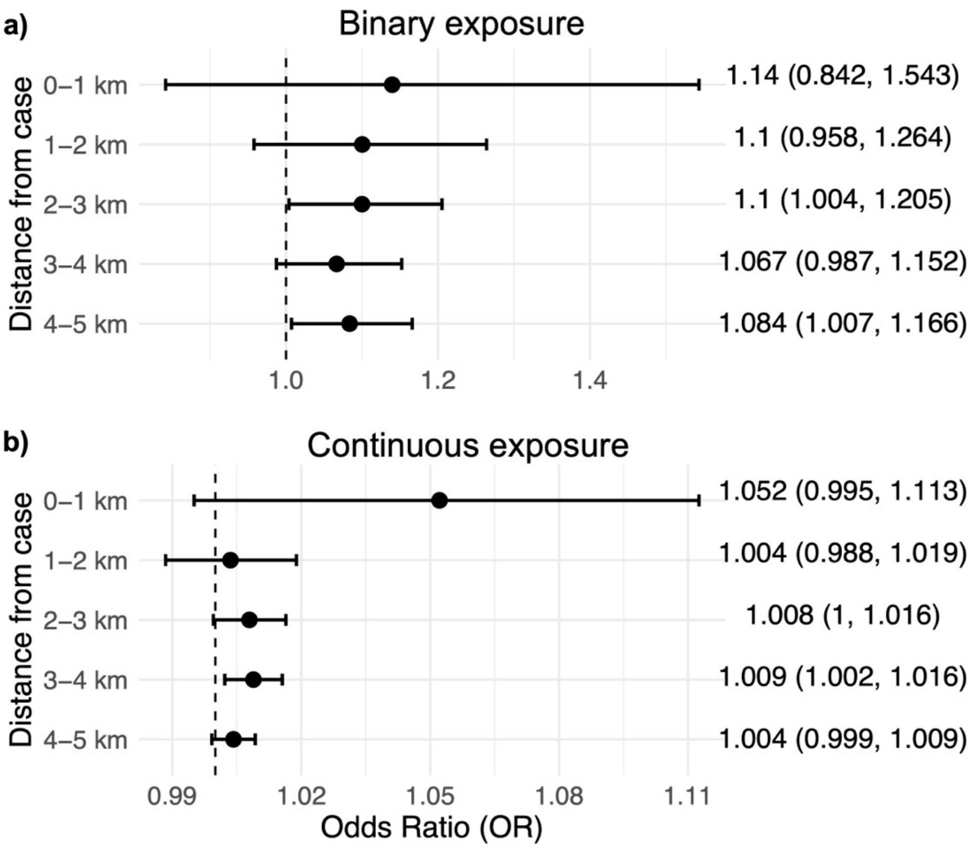


**Figure S7.** Odds ratios (OR) representing the odds of coccidioidomycosis given exposure to oil and gas well construction at varying annuli from the patient residence. Odds ratios are shown for the (a) binary and (b) continuous exposure models. Here, ORs were estimated using a single conditional logistic regression with either the presence/absence (binary) or number (continuous) of wells in each annulus as a predictor.

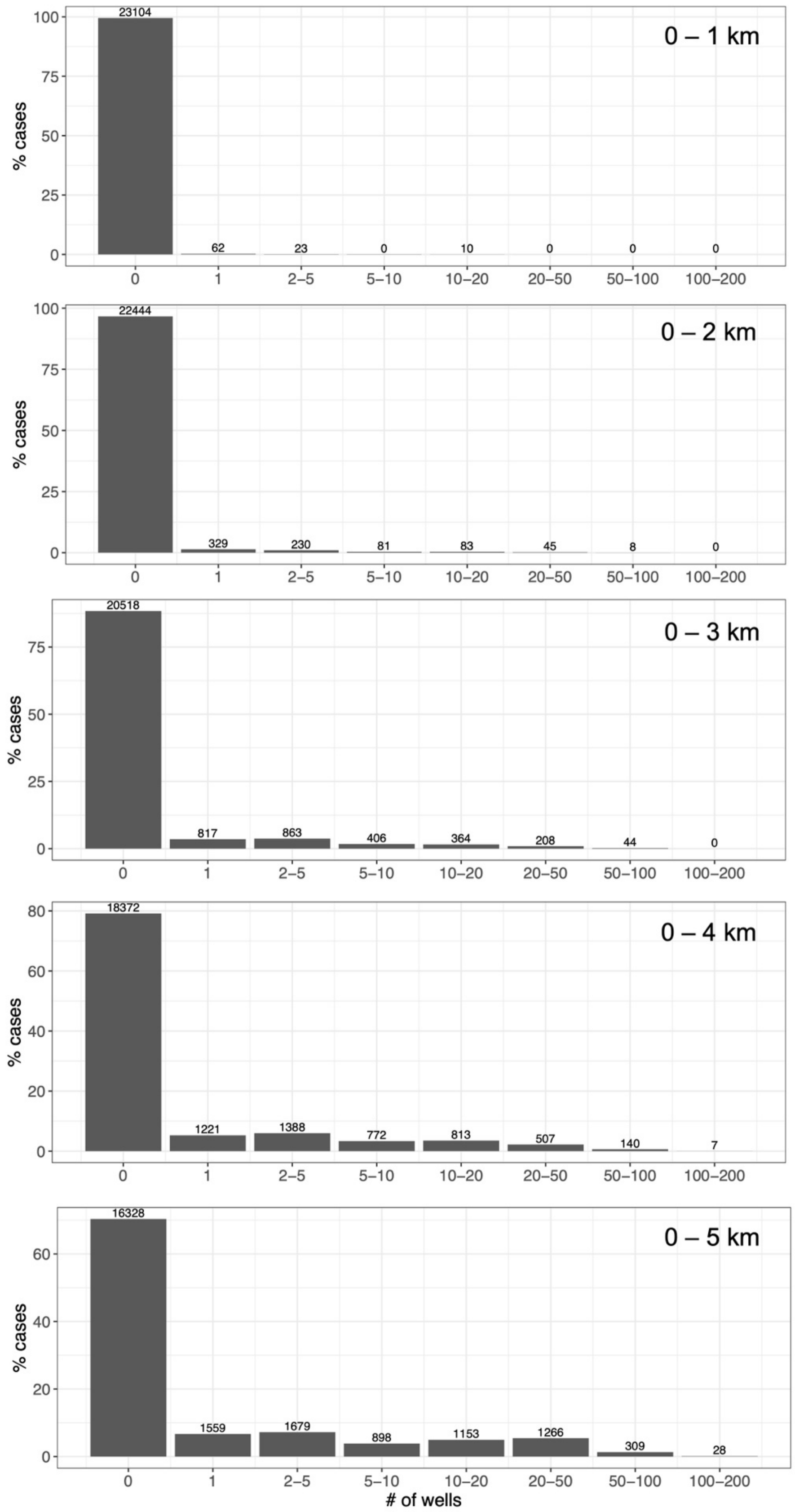


**Figure S8.** Percent of cases with varying levels of well exposure at each distance.

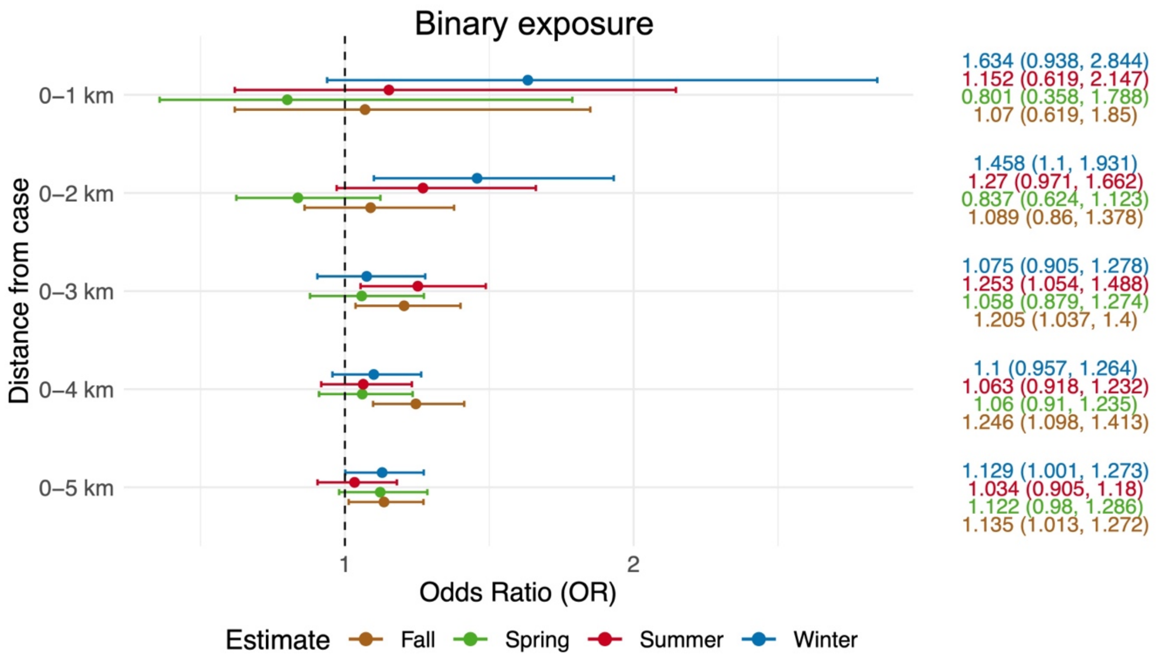


**Figure S9.** Odds ratios (OR) representing the odds of coccidioidomycosis given exposure to oil and gas well construction at varying distances from the patient residence, stratified by season. Odds ratios are shown for the binary exposure model (see Figure 4 for results from the continuous exposure model). The quartile exposure model was not used here as there were not sufficient data within each strata for robust estimation.

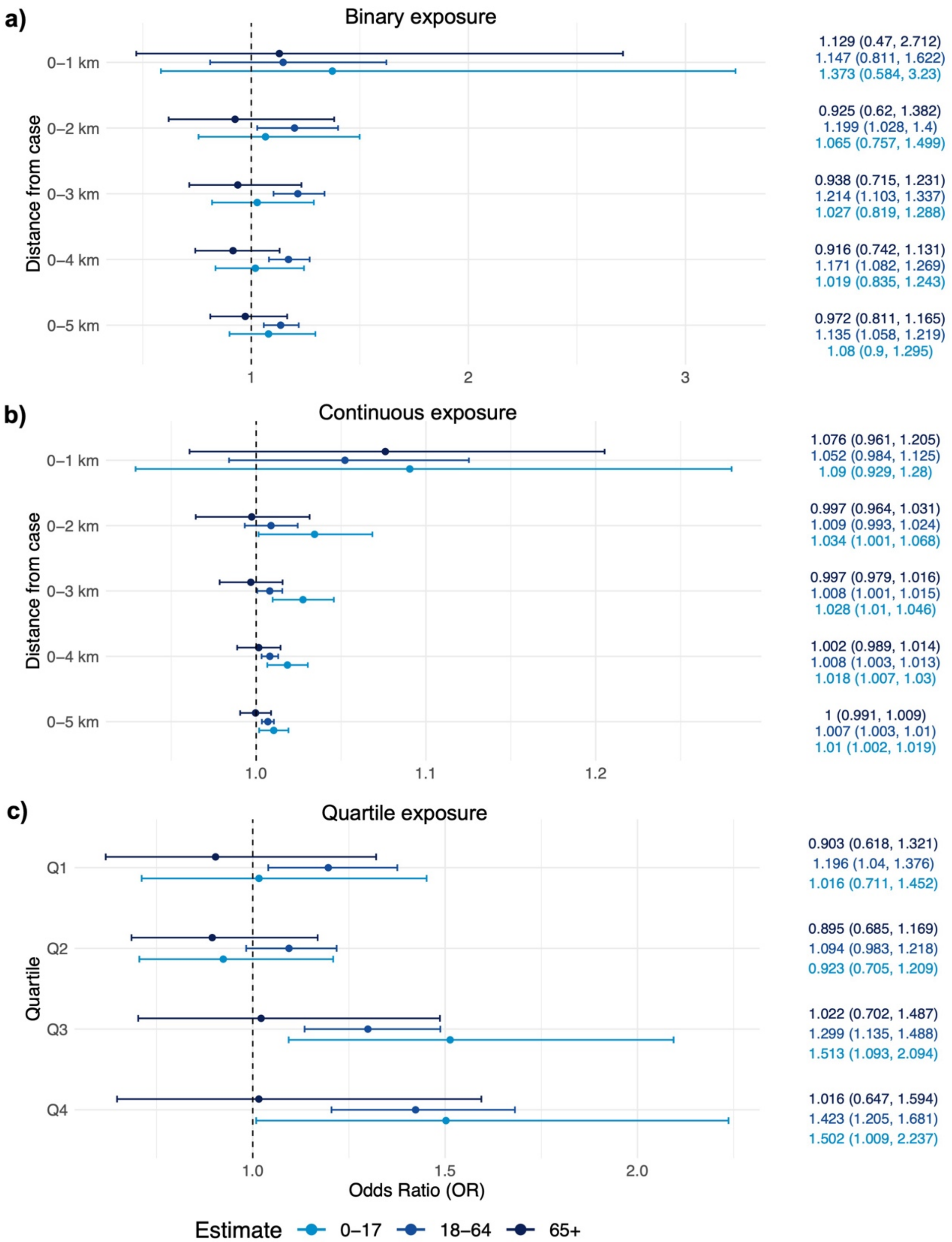


**Figure S10.** Odds ratios (OR) representing the odds of coccidioidomycosis given exposure to oil and gas well construction at varying distances from the patient residence, stratified by age (0-17, 18-64, and 65+). Odds ratios are shown for the a) binary, b) continuous, and c) quartile exposure models. For the quartile exposure, we focus on exposures occurring between 0-5 km and considered only non-zero exposures when calculating quartiles (see *Methods: Exposure Assessment*). Note that the values of the quartiles differ slightly between age groups.

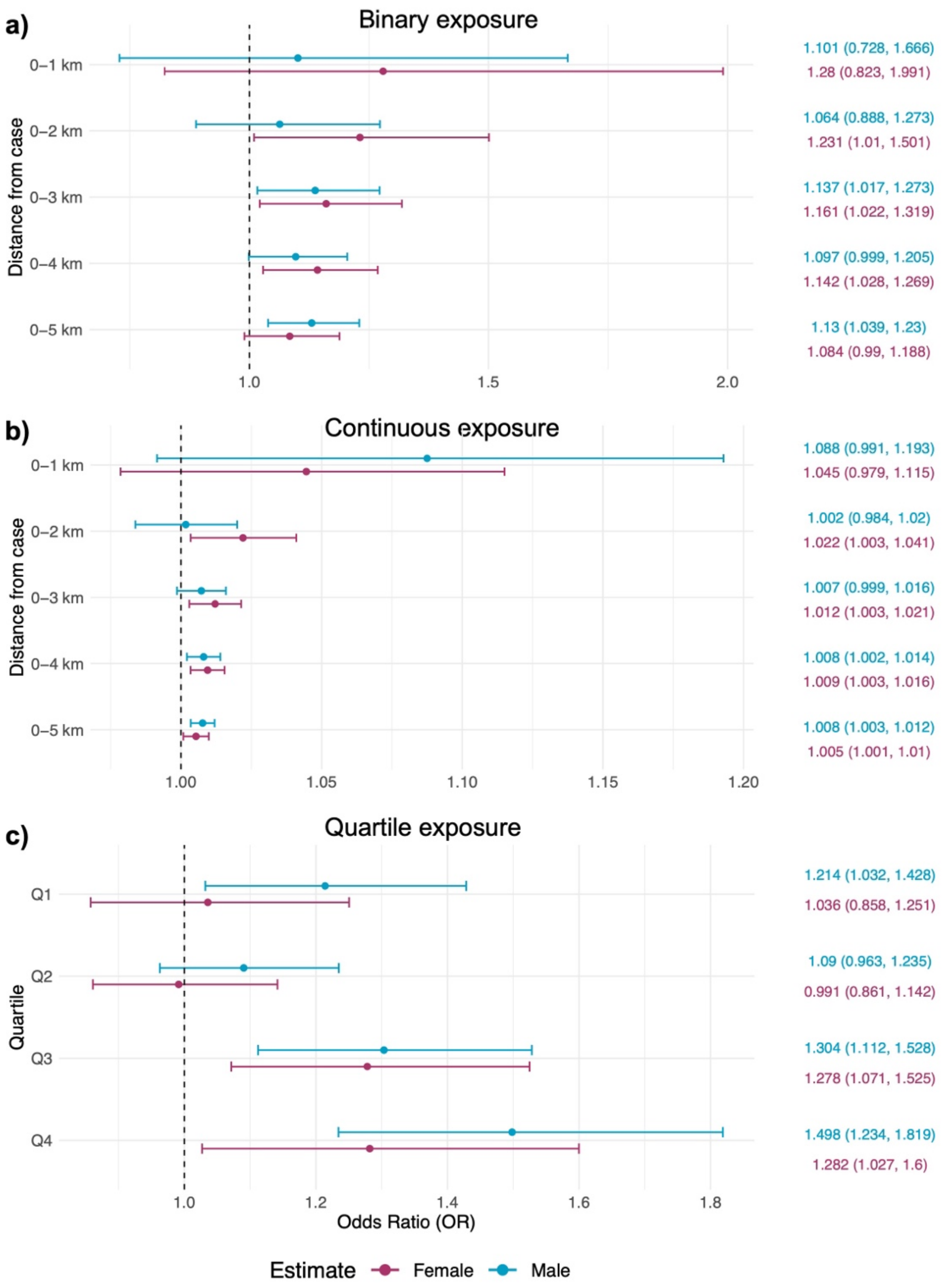


**Figure S11.** Odds ratios (OR) representing the odds of coccidioidomycosis given exposure to oil and gas well construction at varying distances from the patient residence, stratified by sex (male, female). Odds ratios are shown for the a) binary, b) continuous, and c) quartile exposure models. For the quartile exposure, we focus on exposures occurring between 0-5 km and considered only non-zero exposures when calculating quartiles (see *Methods: Exposure Assessment*). Note that the values of the quartiles differ slightly between groups.

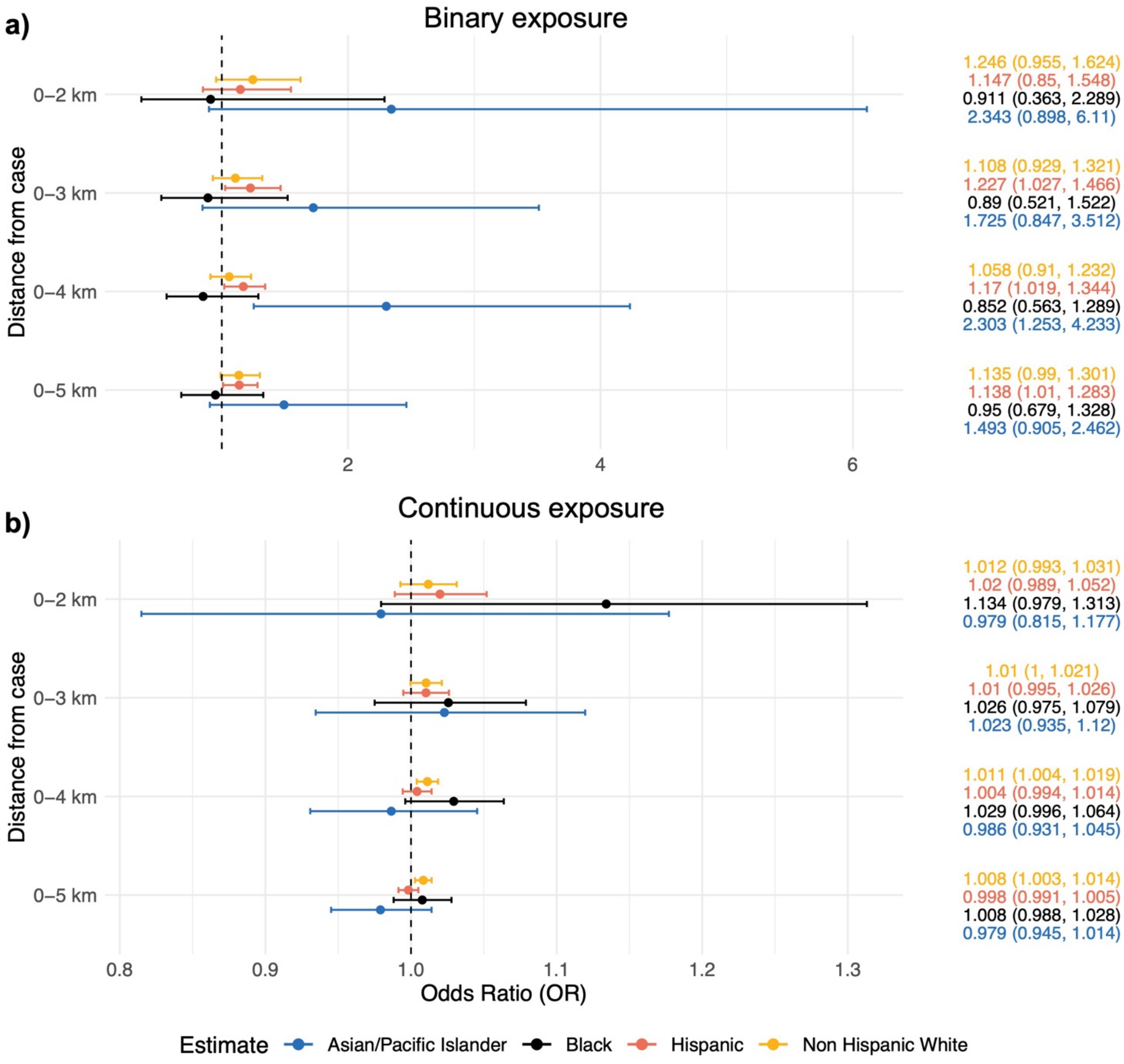


**Figure S12.** Odds ratios (OR) representing the odds of coccidioidomycosis given exposure to oil and gas well construction at varying distances from the patient residence, stratified by race/ethnicity (Asian/Pacific Islander, Black, Hispanic, and Non-Hispanic White). Odds ratios are shown for the a) binary and b) continuous models. The quartile exposure model was not used here as there were not sufficient data within each strata for robust estimation. Similarly, the 0-1 km distance was not included here given the insufficient numbers of certain racial/ethnic groups in this exposure class.
